# L-Dopa and STN-DBS modulate E/I balance and the neural encoding of rhythmic auditory stimulation in Parkinson’s

**DOI:** 10.1101/2025.07.08.25331032

**Authors:** A Criscuolo, M Schwartze, J Schwarz, K Strecker, S Hesse, SA Kotz

## Abstract

Temporally regular auditory stimuli, such as metronome beats or music, are typically utilized in rhythmic auditory stimulation (RAS) to initiate and stabilize the precise temporal coordination of motor plans in persons with Parkinson’s disease (pwPD). Research suggests that RAS promotes the recruitment of the cerebellar-prefrontal network and recalibrates abnormal β-band synchronization in the striato-thalamo-cortical pathway. As these effects resemble those observed via dopaminergic treatments (e.g., levodopa) and deep brain stimulation (DBS) targeting the subthalamic nucleus (STN), one may ask whether RAS may provide a more ecological means to regulate sensorimotor functions in pwPD, and whether treatment combinations may maximize intervention efficacy.

We investigated the influence of both levodopa administration and STN-DBS on the neural encoding of simple isochronous auditory streams in pwPD. A comprehensive analysis of EEG data recorded during temporally regular stimulation revealed changes in (i) β-band event-locked neural responses, (ii) as well as event-related potentials, (iii) neural tracking of rhythm (δ-band inter-trial phase coherence), and (iv) excitation / inhibition balance (E/I; aperiodic exponent) as a function of these treatments. Furthermore, we characterize the link between changes in E/I balance and motor symptom severity (UPDRS-III) with levodopa administration.

Overall, we show inter-individual variability and differential effects of levodopa, 8-week and 1-year after DBS placement on the neural encoding of temporally predictable sounds. Characterizing individual responses to specific treatments is a fundamental step in tailoring rehabilitation protocols and optimizing intervention efficacy.

**Highlights:** In this study we demonstrate that levodopa and DBS interventions differently modulate E/I balance in the brain and how individuals with Parkinson’s process temporally regular auditory streams, as typically provided in rhythmic auditory stimulation protocols. We conclude that characterizing individual treatment response is fundamental to optimize intervention efficacy.

## 1. Introduction

‘One, two, one, two, …’: rhythmic auditory stimulation (RAS) is a widely used non-pharmacological subsidiary intervention to improve gait and balance control in Parkinson’s disease (PD; ^1–7^). Temporally-regular auditory stimuli, such as metronome beats or beats in music, have been used since at least the ‘70s ^1^ to improve motor initiation, coordination, and gait parameters (e.g., velocity, stride length) but also non-motor cognitive functions ^8–10^. Its efficacy has been associated with the strengthening of audio-motor functional connectivity ^11^ and compensatory mechanisms that allow people with PD (pwPD) to switch from an altered internal pacing system (IP; ^12^) to an external cueing system (EC; ^13^). In fact, dopaminergic imbalance in pwPD leads to difficulties in regulating the direct (striato-thalamo-cortico; ^14^) and hyperdirect (subthalamic nucleus (STN) – motor cortex; ^15–17^) pathways, thus impacting action selection and initiation ^18^. RAS can modulate aberrant levels of striatal dopamine observed in pwPD, regulating the cross-talk between the basal ganglia (BG) and the supplementary motor area (direct pathway; ^5,13,14^) and suppressing inhibitory activity in the hyperdirect pathway ^19^. Furthermore, RAS is thought to promote the compensatory recruitment of the EC system, engaging the cerebellar-prefrontal network – initially intact in pwPD – thus recalibrating motor timing and coordination ^4,20^. For example, hypoactivation of BG-cortical and cerebellar-cortical pathways in paced cuing tasks in pwPD is modulated by externally-cued tasks ^13^. Moreover, pwPD who undergo RAS display changes in functional connectivity between the cerebellum, pre-motor cortex, sensorimotor areas, and temporal cortices—even during rest ^21^. This suggests potential long-term benefits of RSA training in gait rehabilitation.

### Factors inducing a shift from the IP to the EC system

The compensatory recruitment hypothesis leverages on the role of *entrainment* to offset the dyscunctional IP system. That is, temporally predictable sensory input provides a grid to precisely time and coordinate body movements ^4,12,22,23^ by modulating underlying event-locked neural dynamics originating in the BG ^16,17,24–26^. Particularly important is the high-frequency activity observed in the STN of the BG system due to hyperpolarization ^27^ driven by dopaminergic degeneration in the substantia nigra ^28–30^. In pwPD, the STN produces aberrant beta-band (12-30Hz; β) activity, leading to increased coherence within the STN and between the STN-globus pallidus (GPi) and STN-motor cortices at rest ^31–33^. PwPD further show altered event-locked β-activity during movement ^34–39^ and listening ^40–42^ and an association between β-activity and symptom severity ^29,35,36,43,44^.

Event-locked β-band dynamics entail an anticipatory (pre-movement and pre-stimulus) as well as event-related (post-movement and post-stimulus) modulations, thought to prepare the sensorimotor system to optimize motor coordination and information processing via predictions ^35,45–48^. In pwPD, atypical β-activity can be compensated by dopamine replacement therapy (levodopa; ^31,32,36,43,49–53)^, which re-calibrates the functional connectivity within the sensorimotor and striato-thalamo-cortical networks ^13,32,43,54,55^ and improves motor symptoms ^32,36,49,50,56^. Similar effects have been achieved via high-frequency deep brain stimulation (DBS) targeting the STN that modulates pathological patterns of hyper-synchronized β oscillations in the striato-thalamo-cortical pathway and improves motor symptoms ^37,57–63^.

These observations reveal a tight link between dopamine, β-band oscillations, and PD symptomatology. Moreover, they suggest that combined interventions (e.g., levodopa + RAS) could optimize clinical outcomes. However, many studies report large inter-individual variability in intervention efficacy ^1,57,64,65^, and possible detrimental effects of levodopa and DBS interventions on cognition (e.g., learning, working memory, cognitive control; ^66–68)^. These findings necessitate a better understanding of the neurofunctional changes associated with traditional interventions at the individual level, and the development of evidence-based, tailored rehabilitation protocols ^69–71^ to better assess the link between treatments (and combinations thereof) and changes in symptomatology. They also demand investigation of the influence of classic interventions on the core mechanism underlying RAS, i.e., the capacity to encode and utilize a temporally regular stimulus train to optimize motor control.Consequently, questions arise as to whether everyone responds to levodopa / DBS interventions the same way and benefits from RAS.

Here, we investigated *if* and *how* levodopa administration and STN-DBS influence the neural encoding of a basic metronome beats in pwPD, as typically provided by RAS. Comprehensive analyses revealed changes in (i) β-band event-locked neural responses, (ii) event-related potentials, (iii) neural tracking of rhythm (delta-band inter-trial phase coherence), and (iv) excitation / inhibition balance (E/I; spectral parametrization) as a function of treatment type. Furthermore, we characterized the link between changes in E/I balance and motor symptom severity (UPDRS-III) with levodopa administration. Overall findings revealed inter-individual variability and differential effects of levodopa, 8-week and 1-year post-DBS on the neural encoding of temporally predictable sound sequences. These results indicate that a better characterization of individual treatment responses, and their link to symptom severity, is necessary to optimize intervention efficacy.

## 2. Materials & Methods

### 2.1. Participants

A total of 13 pwPD (6 females) participated in this study (mean age: 68y, range 40-73y)). On average, the onset age of PD was 49y (± 9.6). All pwPD were right-handed, reported no hearing difficulties, did not use any hearing aids, and reported no additional neurological nor psychiatric comorbidity. Demographic data are provided in Tab. 1.

**Table 1.**
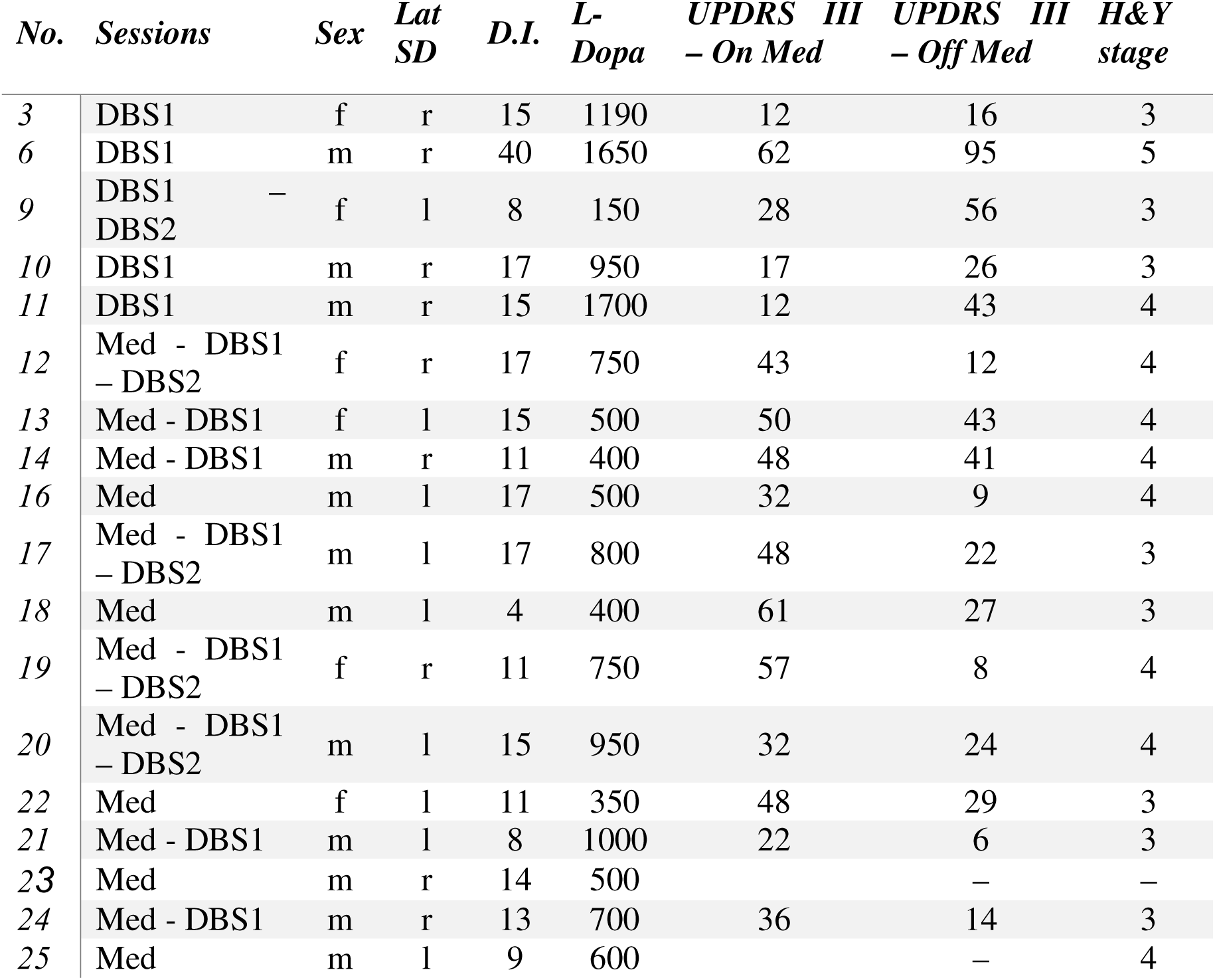
Patients’ demographic data. *Abbreviations: Lat SD = lateralized symptom dominance; D.I. = duration of illness at the moment of the study; L-Dopa = dosage of L-Dopa in mg;, UPDRS III = score on UPDRS part III while on medication and pre-surgery; H&Y stage = disease stage on the Hoehn & Yahr scale pre-surgery*.

Thirteen pwPD took part in the first experimental session ON-/OFF-medication (Med; pre-surgery); of these, 8 took part in the second experimental session, ON-/OFF-DBS (DBS1; 8 weeks post-surgery), along with 5 additional patients; finally, 4 pwPD from the Med session took part in the third part of the experiment, ON-/OFF-DBS (DBS2; 1-year follow-up), along with 1 additional patient. In every session, participants listened to the same temporally regular tone sequences while we recorded their EEG. In the OFF-Med condition, participants were invited to the lab after a 12h washout period.

Participants were recruited and tested at the Max-Planck Institute for Human Cognitive and Brain Sciences in Leipzig, Germany. All participants gave their written informed consent. The study was approved by the local ethics committee at the medical department of the University of Leipzig (Ethics agreement # 225-11-11072011). Participants received 8€/h for taking part in the study.

### 2.2. Clinical characteristics

An experienced neurologist administered the UPDRS (Unified Parkinson’s Disease Rating Scale; (Fahn, Elton, & UPDRS program members, 1987) and the Hoehn and Yahr test (staging of PD. Next to these scales, we collected information regarding the Levodopa replacement therapy (in mg) and lateralized motor symptoms at the onset of PD (left or right).

### 2.3. EEG experiment: design and procedure

#### Temporally regular tone sequence

Participants listened to a 7min long temporally regular tone sequence comprising frequent standard (F0 = 600Hz, duration = 200ms, rise and fall times = 10ms; amplitude = 70dB SPL; STD: 80%) and infrequent deviant (F0 = 660Hz; DEV: 20%) tones. DEV tones differed in pitch from the STD, while all other parameters were kept constant. The tone sequence included a total of 500 tones that were presented continuously and binaurally via two loudspeakers with an inter-stimulus interval of 1.2s, resulting in a stimulation frequency (*Sf*) of ∼.8Hz. Participants were seated in a dimly lit soundproof chamber, and were asked to silently count the deviant tones while they kept their eyes fixated to a white cross in the middle of a computer screen placed on a table in front of them.

As we aimed at assessing neural signatures of basic temporal processing underlying RAS, all subsequent analyses are restricted to standard tones only. We utilized the deviant counting task to merely direct participants’ attention towards the sequence.

### 2.4. EEG recording

The EEG was recorded from 22 Ag/AgCl electrodes mounted in an elastic cap (Electro-Cap International, Inc.) connected to an amplifier (TMS International). These included the scalp locations F7, F3, FZ, F4, F8, CZ, CP5, CP6, P7, P3, PZ, P4, P8, O1, O2 (according to nomenclature of the American Electroencephalographic Society, 1991; i.e. the 10-20 international system). Additional electrodes were placed at both mastoids, A1 and A2. The sternum electrode served as ground. Bipolar electrooculograms (EOGs) were recorded from the lateral canthi (outer corners of the eye) for the horizontal EOG and above and below the right eye for the vertical EOG to control for eye movement and blinks offline. The left mastoid (A1) served as the on-line reference. EEG signals were sampled online at 250 Hz (DC to 40 Hz). Electrode impedances were kept below 3 kΩ.

### 2.5 Data Analysis

#### EEG Preprocessing

EEG data were analyzed in MATLAB, combining custom scripts and the FieldTrip toolbox ^73^. Data were band-pass filtered with a 4th order Butterworth filter in the frequency range of 0.1-50Hz (*ft_preprocessing*). Eye-blinks and other artifacts were identified using independent component analysis. Components with a strong correlation (>.4) with the EOG time-courses were automatically identified and removed with ‘*ft_rejectcomponent*’ before reconstructing the EEG time-course. Next, components were visually inspected to ensure removal of blinks and cardiac artifacts. On average, 2 components were removed (‘*ft_rejectcomponent*’) before reconstructing the EEG time-series. Then we employed artifact subspace reconstruction ( ‘*pop_clean_rawdata*’ function in EEGlab) and an artifact suppression procedure ^74–77^. The artifact suppression procedure interpolated noisy (>100uV) time-windows on a channel-by-channel basis. Lastly, data were low-pass filtered at 40Hz via ‘*ft_preprocessing*’ and segmented to each standard tone onset (−2 to +2s relative to each onset).

#### Time-frequency analyses

After preprocessing, tone-locked EEG data underwent time-frequency transformation (‘*ft_freqanalysis’*) by means of a wavelet-transform ^78^. The bandwidth of interest was set between 1-30Hz, using a frequency resolution of 1Hz. The number of fitted cycles was set to 3 for lower frequencies (up to 4Hz) and to 7 for higher frequencies (above 4Hz). No averaging over channels, trials, or participants was performed at this stage. Data were then re-segmented from -.5 to .5s relative to tone onset and normalized (z-scored).

#### Statistical comparisons

We statistically assessed within-subject (ON-OFF) changes in event-related beta-band (12-25Hz) neural responses by employing Montecarlo permutation testing (‘ft_freqstatistics’) using a two-sided corrected-alpha level of .025 and 500 permutations. The obtained probability map was thresholded at the significance level to obtain a binary mask (logical 0 and 1) which was used to mask out non-significant differences (see Fig. 1). Next, we generated topographic maps of significant pre- and post-stimulus beta-band changes.

**Figure 1.**
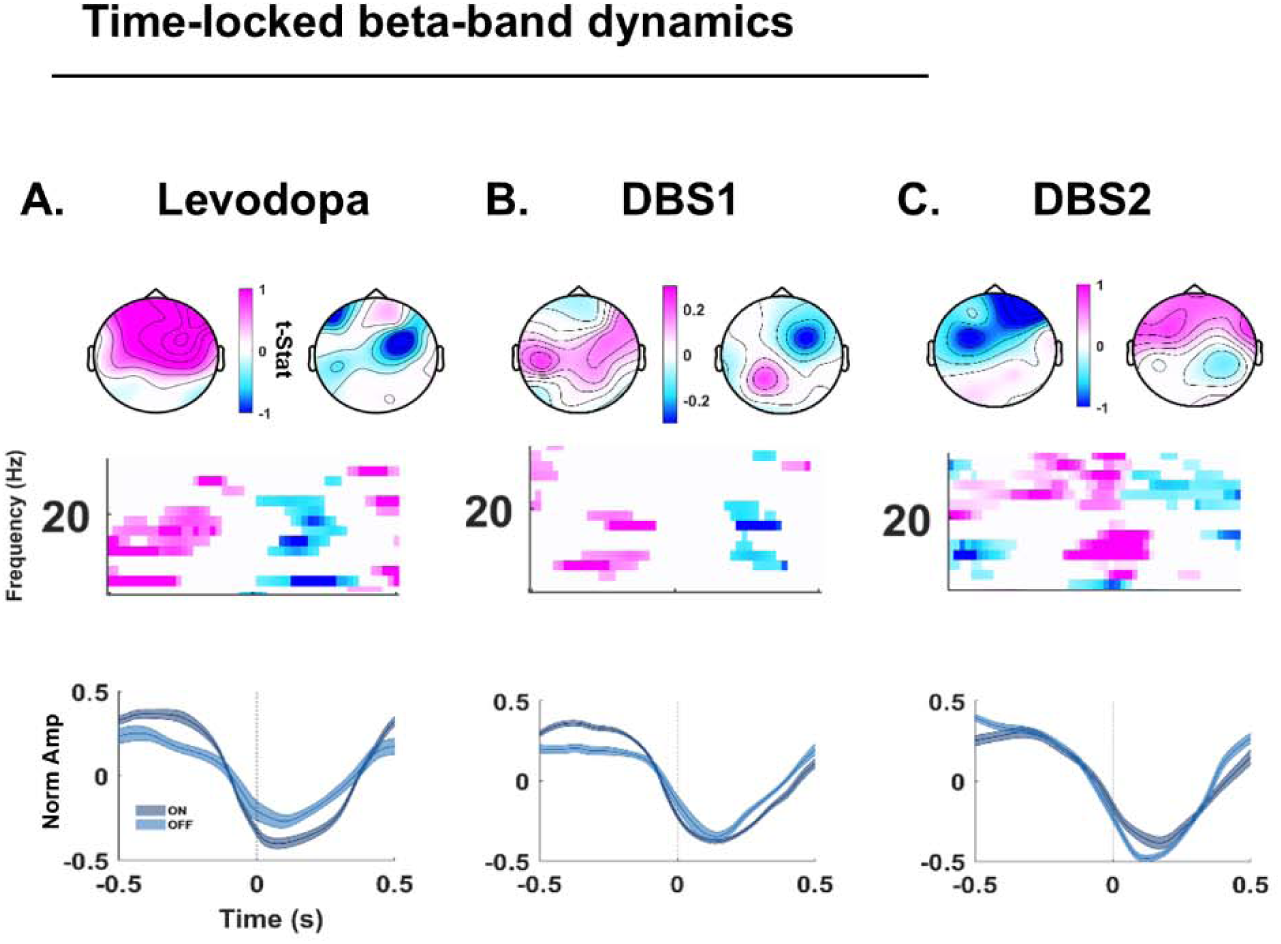
Time-frequency analyses of beta-band dynamics. *A-C: Figure displays time-locked beta-band (12-25Hz) power changes in the Levodopa (A), DBS1 (B; 8-week follow-up) and DBS2 (C; 1-year follow-up) sessions by comparing the ON – OFF conditions. On top, the topographic maps of significant beta-band activity in pre-and post-stimulus interval respectively. In pink, positive t-values; blue for negative t-values. Color-coding and t-values in the color bar are used for both the topographic maps and the spectrograms underneath. The corresponding spectrograms are provided in the middle: time on the x-axis (-.5 to .5s relative to sound onset) and frequency on the y-axis (12-25Hz). The plots are masked by the probability maps obtained by Montecarlo permutation testing. Non-significant within-subject effects are, thus, masked out (white). At the bottom, beta-activity in the ON (dark blue) and OFF (light blue) condition. Solid lines report the group mean, while the shades represent the standard error across participants, after averaging across channels and trials*.

#### Time-series plots for group differences

To better visualize event-locked beta-band temporal dynamics, we created time-series plots (Fig. 1, right). These plots show the group grand-average for each condition (solid line) and the standard error across participants (shades).

#### ERP analyses

We assessed within-subject changes in the amplitude, latency, and variability of event-related potentials (ERP) to standard tone onsets. Preprocessed data were low-pass filtered at 30Hz via ‘*ft_preprocessing*’ and averaged across a fronto-central cluster of channels (FC); ’F3’, ’F4’, ’F7’, ’F8’,’FC3’, ’FC4’), thus excluding parietal and posterior electrodes.

Single-trial event-related responses to standard tones were re-segmented (-.8 to .6s relative to tone onset). To perform peak analyses, we identified, at the single participant and trial level, the P50, N100, and P200 amplitude and latency. We created windows of interest centered at 50, 100 and 200ms respectively, and extending 30ms before and after (e.g., for the P50, we used a window ranging from 20 to 80ms relative to tone onset). Thus, we calculated the mean amplitude and peak amplitude latency in each time window. ERP morphology results are provided in Fig. 2A.

**Figure 2.**
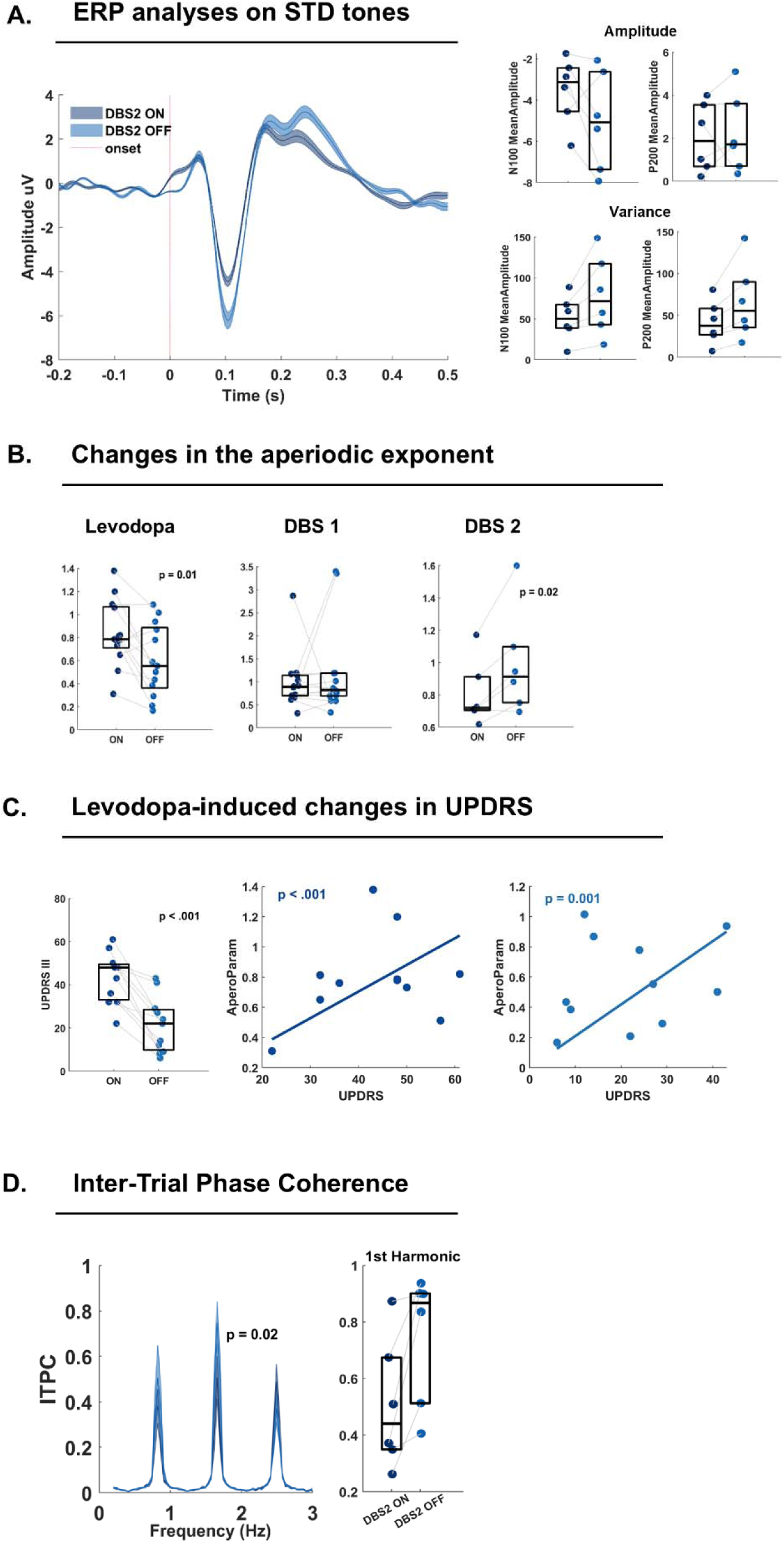
ERP, spectral parametrization, UPDRS and ITPC. A. *Event-related potentials (ERP) for DBS2 (1-year follow-up) ON (dark blue) and OFF (light blue) stimulation. Solid lines represent the group mean, while the shades report the standard error across participants after averaging across the FC cluster of interest and over trials. The red dotted vertical line represents the STD tone onset. Time on the x-axis (-.2 to .5s) and Amplitudes in uV on the y-axis. On the right side, ERP peak analyses for the N100 and P200 mean amplitudes (top) and the variance across trials per participant (bottom). Color coding as in the ERP; one dot per participant. The central horizontal line in each boxplot represents the median, while the edges of the box represent the 25th and 75th percentiles. B: boxplots representing changes in the aperiodic exponent in function of ON-OFF sessions, per condition (Levodopa on the left; DBS1 in the middle; DBS2 on the right). C: on the left, levodopa-induced changes in the UPDRS-III. On the right, the linear regression assessing the link between individual aperiodic exponent and UPDRS-III scores in the ON (middle plot) and OFF (right plot) conditions. D: results from the inter-trial phase coherence (ITPC) analyses. On the left, the ITPC frequency spectrum: frequency (.2-3Hz) on the x-axis and coherence on the y-axis. On its right, individual coherence peaks in the first harmonic of the stimulation frequency (1.6Hz) color coded per condition (dark blue for DBS2-ON and light blue for DBS2-OFF)*.

##### Mixed Effect Models for ERP data

We assessed differences in the morphology of P50, N100, P200, and P300 via Mixed effect models (‘*fitlme*’ function in MATLAB, version 2024). The model included ‘Session’ as within-subject factor, and a random intercept per participant. We fitted separate models for each experimental condition: ON-OFF (i) Med, (ii) DBS1 and (iii) DBS2.

#### Spectral parametrization

To investigate levodopa-and DBS-induced changes in the excitation/inhibition (E/I) balance across conditions, we performed spectral parametrization analyses ^79,80^. Differently from typical Fourier (FFT) analyses, this approach parametrizes data into a periodic and an aperiodic component (i.e., the 1/F typically observed in the Fourier spectra; ^81^). Next, the slope of the aperiodic component (the exponent of the decay function) was utilized as a proxy for E/I.

Continuous pre-processed data underwent the automated spectral parameterization algorithm described in ^81^ and implemented in FieldTrip in a two-step approach. First, the power spectral density was estimated using the multi-taper method based on the Slepian sequence (‘ft_freqanalysis’ was used in combination with the multi-taper method for FFT (‘mtmfft’) and power as output (‘pow’)), and secondly, by specifying ‘fooof_aperiodic’ as output. The output frequency resolution was set at 0.2Hz.

##### Statistical analyses

Subsequent statistical analyses were performed on the same FC cluster as described above. Differences in E/I balance across conditions (ON-OFF per treatment) were statistically assessed by comparing the aperiodic exponent via paired-sample t-test. A *p-*value below .05 was considered statistically significant.

#### ITPC

We used inter-trial phase coherence (ITPC) analyses to assess whether levodopa and DBS interventions modulated the neural encoding of temporally predictable tone sequences. The complex FFT spectrum was obtained by performing FFT decomposition at the single-participant and -channel level into 20s-long moving time segments along the entire tone sequence. Next, the ITPC spectrum was calculated by dividing the Fourier coefficients by their absolute values (thus, normalizing the values to be on the unit circle), calculating the mean, and finally taking the absolute value of the complex mean. Further documentation can be found on the FieldTrip website (https://www.fieldtriptoolbox.org/faq/itc/). For illustration purposes, the ITPC spectrum was restricted to .2-3Hz.

#### Statistical comparisons

Statistical analyses were performed on the same FC cluster as described above, and by focusing on two frequencies of interest: the stimulation frequency (.8Hz) and its first harmonic (1.6Hz), as typically done in previous EEG studies investigating rhythm processing (e.g., ^82^). Single-participant data were compared across sessions via paired-sample t-test. A *p-*value below .05 was considered statistically significant.

### 2.5. Data and code Availability

The analysis code in use will be provided as open access resource. Lesion data and participant data in use are under data protection rules.

## 3. Results

### 3.1. Time-frequency analyses

Via time-frequency analyses, we assessed the modulatory effects of Med (Levodopa) and DBS 1-2 on beta-band (12-25Hz) event-related responses to STD tones (Fig. 1).

In Med (Fig. 1A) as well as in DBS1 (Fig. 1B), pre-stimulus (-.5 to -.3s) low-beta (12-20Hz) power was larger in the ON-as compared to the OFF-session, and became more negative post-stimulus (∼.1 to .3s). In both cases, the pre-stimulus effect was prominent over fronto-central electrodes, while the post-stimulus effect was mostly right-lateralized. In the DBS2 condition, we found the same effect, but in the upper (20-25Hz) beta-band (Fig. 1C). In the low-beta range, there was only a post-stimulus (∼0 to .2s) effect. Topographic plots showed a prominent fronto-central distribution of activity, bilaterally.

### 3.2. ERP peak analyses

We performed ERP morphology analyses to assess the effect of Med and DBS via a linear mixed model effect (LME). We employed separate LME to assess the amplitude and latency of P50, N100 and P200 components of the ERPs.

LME only revealed an effect of DBS2 on the N100 mean amplitude (*p* = .025; Tab. 2; Fig. 2A). In particular, the N100 was increased when DBS2 was ON as compared to when it was OFF, mirroring results observed in the post-stimulus low-beta activity (Fig. 1C).

**Table 2.**
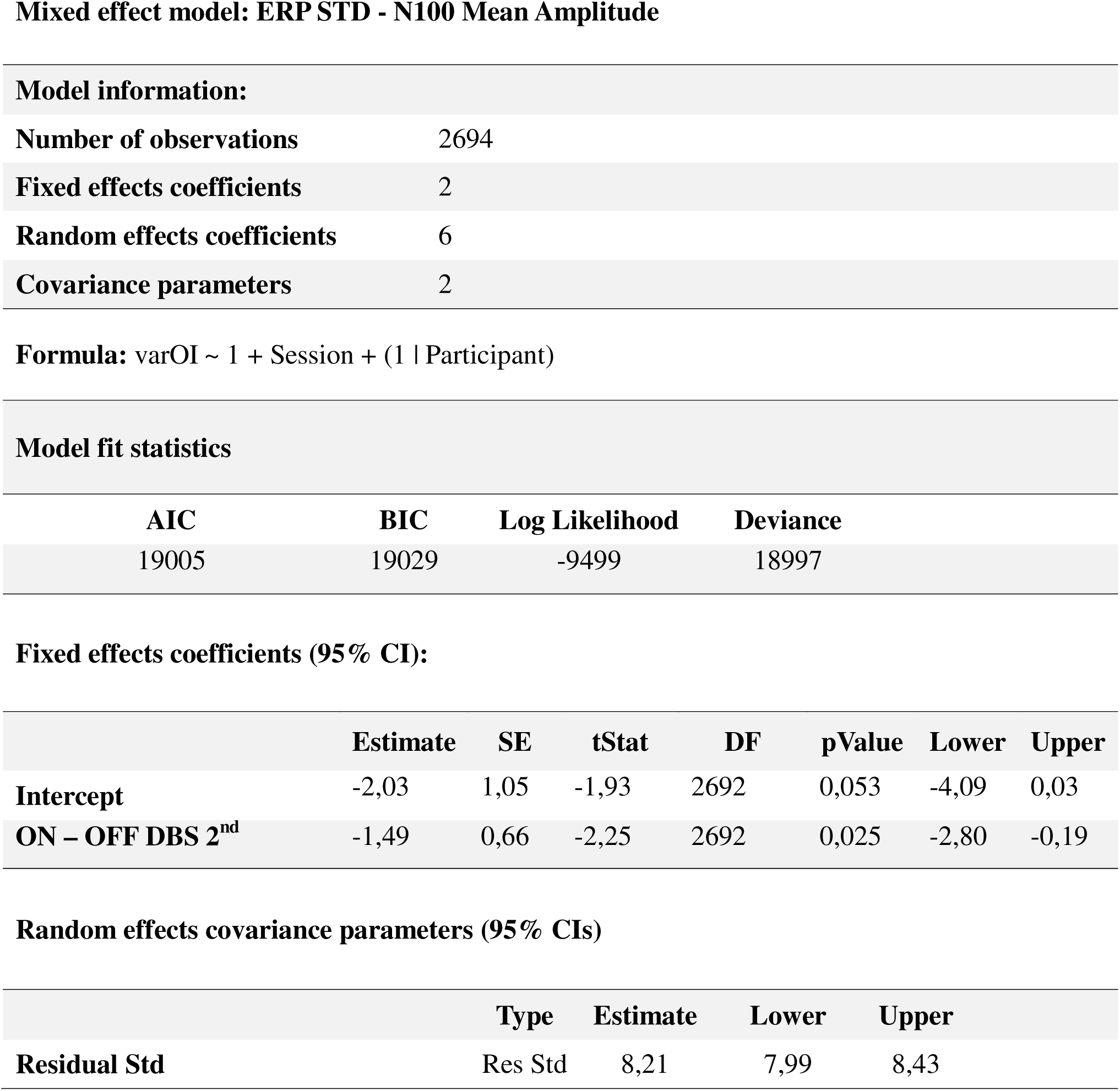
Mixed effect model on ERP N100 Mean Amplitude in DBS2 ON-OFF. *The table reports model information: number of observations, fixed effect coefficients, random effect coefficients, covariance parameters. Then, the formula used to fit the model and model fit statistics: AIC, BIC values, Log Likelihood and Deviance. Further below, the fixed effect coefficients in a 95% confidence interval (CI): estimate, standard error, t-stat, degrees of freedom (DF), p value, lower and upper bound. Right below, random effects covariance parameters: estimate, lower and upper bound*.

Model fit statistics
| AIC | BIC | Log Likelihood | Deviance |
| --- | --- | --- | --- |
| 19005 | 19029 | -9499 | 18997 |

Fixed effects coefficients (95% CI):
|  | Estimate | SE | tStat | DF | pValue | Lower | Upper |
| --- | --- | --- | --- | --- | --- | --- | --- |
| Intercept | -2,03 | 1,05 | -1,93 | 2692 | 0,053 | -4,09 | 0,03 |
| ON – OFF DBS 2<sup>nd</sup> | -1,49 | 0,66 | -2,25 | 2692 | 0,025 | -2,80 | -0,19 |

Table 2 - Mixed effect model on ERP N100 Mean Amplitude in DBS2 ON-OFF.
|  | Type | Estimate | Lower | Upper |
| --- | --- | --- | --- | --- |
| Residual Std | Res Std | 8,21 | 7,99 | 8,43 |

### Mixed effect model: ERP STD - N100 Mean Amplitude

### 3.3. Spectral parametrization

After parametrizing the Fourier spectrum into a periodic and aperiodic components, we statistically assessed within-subject differences in the exponent of the aperiodic slope via paired-sample t-test. We found an increased exponent in the Med-ON as compared to –OFF condition (*p* = .01; Fig. 2B) and an opposite effect in the DBS2 condition (*p* =.02).

Via linear regression analyses, we further showed a statistically significant positive association between individual aperiodic exponents and UPDRS-III scores in both the ON (*p* < .001; Fig. 2C) and OFF *p* = .001) conditions.

### 3.4. Inter-Trial Phase Coherence

The imaginary part of the complex Fourier spectrum was used to calculate the Inter-trial phase coherence (ITPC). Statistical analyses assessed within-subject differences in ITPC at the stimulation frequency (.8Hz) and its first harmonic (1.2Hz). Paired-sample t-tests only showed an effect of DBS2, where ITPC was higher in the –OFF as compared to the –ON session, but only for the first harmonic (*p* = .02; Fig. 2D).

## 4. Discussion

Parkinson’s disease (PD) is a neurodegenerative disorder whose incidence has more than doubled in the past 30 years ^83^. Dopaminergic changes in the striatum lead to movement disorders ^29,84^ and significantly impact patient’s daily routines and quality of life. As such, PD represents an ever-growing burden for individuals, their families, and the public health systems.

Dopamine replacement therapy (e.g., via levodopa; ^31,32,36,43,49–53)^ and deep brain stimulation (DBS) protocols targeting the sub thalamic nucleus (STN; ^37,57–63)^ are the most common interventions utilized to improve motor coordination and PD symptomatology. PwPD often undergo subsidiary training to improve motor control, such as rhythmic auditory stimulation (RAS). In RAS, temporally predictable sensory input are thought to provide a framework to facilitate the precise coordination of body movements fostering audio-motor connectivity ^4,11,22^.

Although the majority of experimental evidence is quite promising and show beneficial effects of these three intervention types, there is largely unexplored interindividual variability in therapeutical outcomes ^1,57,64,65^. Not only the therapy may not work for everyone, but sometimes levodopa and DBS treatments can impact cognitive functions such as learning, working memory, cognitive control ^66–68^ and even speed up cognitive decline ^67^. The large variability and the increasing incidence of the disease demand for a deeper understanding of *how* and *why* treatments may or may not work. Furthermore, these observations demand for the development of evidence-based, tailored rehabilitation protocols ^69–71^ which thoroughly assess *which* treatment may function *best* for *each* individual.

In this EEG study, we investigated whether levodopa and DBS interventions alter the way pwPD processed rhythmic auditory sounds as typically involved in RAS, and further examined the influence of these interventions on neural signatures of excitation / inhibition (E/I) balance.

First, we focused on event-locked beta-band activity (β ; 12-25Hz). PwPD typically show altered event-locked β activity during movement ^34–39^ and listening ^40–42^. β-band activity is also correlated with symptom severity ^29,35,36,43,44^. Our analyses of event-locked β activity showed similar effects of levodopa and DBS (in the first follow-up 8 weeks from surgery): Levodopa replacement increased pre-stimulus β-band activity and its post-stimulus negativity peak. In the second follow-up (1 year after surgery), we found a dissociation between high-(20-25Hz) and low-β (12-20Hz) activity: while the former mimicked the pre-and post-stimulus effects observed in levodopa and DBS1, low-beta displayed a post-stimulus increase in the peak negativity for the OFF as compared to the ON condition.

The anticipatory pre-stimulus β activity is considered to prepare the sensorimotor system to optimize information processing via predictions ^35,45–48,76,77^. In agreement with previous studies employing levodopa ^31,32,36,43,49–53^ and DBS ^37,57–63^, we confirm that these intervention types can significantly modulate event-locked β dynamics. Particularly, we mirror results reported in ^40^ comparing pwPD with healthy controls, as we showed a reduction in pre-stimulus β power and post-stimulus negativity in pwPD OFF levodopa and DBS1 as compared to the ON state. Authors interpreted these results as reflecting abnormal engagement and disengagement of sensorimotor regions ^40^. These observations also align with other evidence linking levodopa and DBS to increased time-locked β power ^37,53,56^ and the readiness potential ^85,86^, reflecting internalized timing and anticipation of event onsets ^87^. However, contrasting evidence reported reductions of event-related activity ^41,42^ and thus, no improvement in audio-motor entrainment. Our results on DBS2 (1 year follow-up after surgery) also raise doubts on the specific effects of the DBS intervention on β dynamics.

How can we reconcile these apparent discrepancies? Two discussion points deserve further consideration: the link between β, auditory and rhythm processing; and the differentiation between high-and low-β activity.

### Rhythm and temporal processing

Although the basal ganglia (BG) system plays a fundamental role in rhythm and beat processing in both human and nonhuman species ^26,75,77,88,89^, dopaminergic degeneration in the BG system does not necessarily impair rhythm and beat processing in pwPD ^1^. For example, pwPD show preserved capacity to encode temporal information ^40^, but a heterogeneous capacity to display predictive motor timing behavior in the subsecond range ^12,40,90–93^. This observation supports the hypothesis that the BG and dopaminergic systems may not be merely encoding temporal information, but rather play fundamental role in generating temporal predictions and supporting synchronization to rhythm ^1,94–96^. Such views align with the compensatory recruitment hypothesis: RAS may promote the switch from an impaired interval pacing system (engaging direct and hyper-direct pathways) to the external cueing (EC) system (engaging cerebellar-prefrontal network – initially intact in pwPD), thus recalibrating motor timing and coordination^4,20^, potentially by inducing dopaminergic bursts from the ventral tegmental area ^97,98^.

To probe this hypothesis, we employed inter-trial phase coherence (ITPC) analyses to assess whether levodopa and DBS modulated the neural encoding of the temporal regularity in temporally regular tone sequences. We found no temporal encoding differences under dopaminergic and DBS1 (8-week follow-up) treatment, confirming that temporal encoding is intact in pwPD and is not altered by either intervention types. However, in the 1-year follow-up (DBS2), we found that the spectral peak in the first harmonic (1.2Hz) of the stimulation frequency (.8Hz) was reduced in the ON compared to the OFF condition. This may suggest a worsening of temporal encoding in the 1-year follow-up, aligning well with the observations drawn from the low-β analyses in DBS2. So do low-and high-β-band activity convey complementary but distinct information in rhythm processing?

### Differentiation between low- and high-β-band activity

Although typically treated as a homogenous frequency-band, low- and high-β frequencies have been differentiated into motor preparation and temporal prediction, respectively ^99,100^. This refined characterization is supported by clinical evidence showing that PD affects more the low-than the high-β band, and levodopa / DBS can selectively re-balance low-β activity ^51,60,101^. Our results further confirm modulatory effects of levodopa and DBS1 on the low-beta band, aligning with previous evidence and strengthening the notion that PD impairs temporal predictions. In the DBS2 condition, however, the effects observed in the levodopa and DBS1 shifted to the high-β band, while the low-β band displayed increased event-related response in the DBS2-ON condition.

Does long-term DBS modulate how pwPD process basic auditory and temporal information differently from levodopa and short-term DBS? This question motivated follow-up analyses exploring broad-band event-related responses (ERP) and shifts in the excitation (E/I) balance.

### Increased ERP amplitudes and E/I balance in DBS2

We employed ERP morphological analyses to assess the modulatory effects of levodopa and DBS on event-related responses. Statistical analyses showed an increased N100 ERP amplitude in the DBS2 OFF compared to ON condition, thus paralleling the effects observed for low-β. There were no modulatory effects in the levodopa and DBS1 conditions.

In line with the current findings, Gulberti et al., ^102^ reported that while dopaminergic medication and short-term STN-DBS did not modulate ERP amplitude, long-term implantation reduced N100 responses; hence, turning-off DBS resulted in an increased N100 response. Opposing evidence, however, exists showing that STN-DBS increased N100 responses in pwPD ^103^. Increased N100 amplitudes in response to temporally predictable auditory sequences ^74,104–110^ have been associated with hypersensitivity ^74,104^ to sensory input in aging. Namely, aging individuals are thought to not efficiently employ suppression mechanisms to regulate cortical responses to predictable and repeated sensory stimulation ^109,111^. The hypersensitivity hypothesis in aging is further supported by spectral parametrization analyses, showing an increased aperiodic exponent of the 1/F ^74^, signaling an imbalanced E/I ^81,112^.

### Can we draw parallels between these observations in aging and and pwPD?

Motivated by these observation, we investigated whether levodopa and DBS modulated the E/I balance, as measured via spectral parametrization analyses. Utilizing the exponent of the 1/F slope, we showed that while levodopa increased the E/I ratio, DBS2 decreased it. Previous evidence reported that both dopaminergic medication and STN-DBS increased the aperiodic exponent in the STN, and associated the increased E/I to changes in the hyperactivity of the STN^113^. In follow-up analyses, we were further able to show a positive link between the aperiodic exponent and individual UPDRS-III scores: increased aperiodic exponents were associated with worse motor symptoms. This observation strengthens the idea that E/I balance could be a valid biomarker for PD symptomatology ^113^. Complementing this notion, the current DBS-2 results showed that stimulation could reduce aberrant E/I balance: turning off the stimulation, increased the E/I ratio, thus speaking in favor of positive treatment effects of long-term DBS.

### Future outlook

#### Levodopa re-calibrates β activity, but alters the E/I balance

How can we reconcile the levodopa-induced increased E/I (detrimental effect) with the modulatory influence on the temporal encoding of RAS (positive effect)? One may argue that while levodopa re-calibrates β activity ^31,32,36,43,49–53^, it may have detrimental effects on other cognitive functions ^66–68^. This hypothesis demands for further investigations assessing multiple cognitive functions along with electrophysiological data, so to achieve a better understanding of the effects of levodopa therapy on motor control and general cognition.

#### DBS may differentially affect short- and long-term modulatory effects

How can we interpret the apparently divergent findings gathered from DBS-1 and DBS-2? We propose that DBS may lead to differential short- and long-term effects ^102^. While we observed positive effects of DBS-1 on time-locked β activity, we found no modulatory influence on ERPs ^102^, ITPC, and E/I, potentially indicating intact auditory and temporal processing. In DBS-2, however, we observed that switching-off the stimulation altered the E/I balance, and increased event-locked neural responses to fully predictable sounds (ERP and β activity). These observations may indicate a neuroplastic adaptation to the STN stimulation, and a dependence on it for recalibrating altered network connectivity ^37,57–63^. These observations encourage future research to further characterize the long-term effects of DBS on neurocognitive functions.

#### Complementary approaches

While our focus was predominantly on the dopaminergic circuit and β activity, we know that PD is a multifactorial disorder featuring degeneration of the cholinergic and the noradrenergic pathways ^114^, a variegated interaction of motor, cognitive and motivational symptoms ^71^, and alterations in neural activity beyond β, including theta (matching the typical tremor frequency) ^44^ delta ^115^ and alpha rhythms ^40^. The present EEG setup further prevented any characterization of network activity and source localization, which represent fundamental steps to better characterize modulatory interactions within and across sensorimotor and cortico-subcortical pathways. Thus, we encourage future multimodal imaging approaches—possibly combining functional magnetic resonance imaging with magnetic resonance spectroscopy, and EEG—to capture both spatial and temporal dynamics of auditory-motor coupling ^4^, as well as neurotransmitters levels across the brain. Such a comprehensive approach would advance our understanding of the neuropathological changes in PD, and would support the development of multimodal and individualized therapeutical approaches.

## 5. Conclusions

RAS is a subsidiary training that can be applied in parallel with dopaminergic replacement therapy and DBS that are commonly used to improve motor coordination and gait in pwPD. All three of these interventions are thought to re-calibrate altered network connectivity in the direct and hyperdirect pathways, to reduce aberrant beta activity, and to promote a possible switch to an intact cerebello-thalamo-cortical pathway.

However, does everyone benefit from RAS? Do levodopa - DBS treatments influence the way people respond to RAS training? We characterize inter-individual variability and differential effects of levodopa, 8-week and 1-year DBS treatments on the neural encoding of basic sounds and rhythm. In doing so, we aim to encourage future studies to adopt a multimodal and individualized approach: if we want to optimize rehabilitation efficacy, we need to better understand individual neurocognitive responses to treatments.

## Data Availability

Lesion data and participant data in use are under data protection rules.

## Notes

### Competing Interest Statement

The authors have declared no competing interest.

### Funding Statement

The study did not receive any specific funding.

### Author Declarations

Participants were recruited and tested at the Max-Planck Institute for Human Cognitive and Brain Sciences in Leipzig, Germany. All participants gave their written informed consent. The study was approved by the local ethics committee at the medical department of the University of Leipzig (Ethics agreement # 225-11-11072011).

### Summary of Updates

Minor edits in the manuscript (spelling mistakes) and updated authors list with correct affiliations

## References

1. Nombela, C., Hughes, L. E., Owen, A. M. & Grahn, J. A. Into the groove: Can rhythm influence Parkinson’s disease? Neurosci Biobehav Rev 37, 2564–2570 (2013).

2. Burrai, F., Apuzzo, L. & Zanotti, R. Effectiveness of Rhythmic Auditory Stimulation on Gait in Parkinson Disease: A Systematic Review and Meta-analysis. Holist Nurs Pract 38, 109–119 (2024).

3. Braun Janzen, T., Koshimori, Y., Richard, N. M. & Thaut, M. H. Rhythm and Music-Based Interventions in Motor Rehabilitation: Current Evidence and Future Perspectives. Front Hum Neurosci 15, (2022).

4. Koshimori, Y. & Thaut, M. H. Future perspectives on neural mechanisms underlying rhythm and music based neurorehabilitation in Parkinson’s disease. Ageing Res Rev 47, 133–139 (2018).

5. Wang, L. et al. Effects of Rhythmic Auditory Stimulation on Gait and Motor Function in Parkinson’s Disease: A Systematic Review and Meta-Analysis of Clinical Randomized Controlled Studies. Front Neurol 13, 818559 (2022).

6. Bella, S. D., Benoit, C. E., Farrugia, N., Schwartze, M. & Kotz, S. A. Effects of musically cued gait training in Parkinson’s disease: Beyond a motor benefit. Ann N Y Acad Sci 1337, 77–85 (2015).

7. Bella, S. D. et al. Gait improvement via rhythmic stimulation in Parkinson’s disease is linked to rhythmic skills. Sci Rep 7, 1–11 (2017).

8. Benoit, C.-E. et al. Musically Cued Gait-Training Improves Both Perceptual and Motor Timing in Parkinsonâ€^TM^s Disease. Front Hum Neurosci 8, 1–11 (2014).

9. Wang, L. et al. Effects of Rhythmic Auditory Stimulation on Gait and Motor Function in Parkinson’s Disease: A Systematic Review and Meta-Analysis of Clinical Randomized Controlled Studies. Front Neurol 13, 818559 (2022).

10. Thaut, M. H., McIntosh, K. W., McIntosh, G. C. & Hoemberg, V. Auditory rhythmicity enhances movement and speech motor control in patients with Parkinson’s disease. Funct Neurol 16, 163–172 (2001).

11. Thaut, M. H. The Future of Music in Therapy and Medicine. Ann N Y Acad Sci 1060, 303–308 (2005).

12. Jones, C. R. G., Malone, T. J. L., Dirnberger, G., Edwards, M. & Jahanshahi, M. Basal ganglia, dopamine and temporal processing: Performance on three timing tasks on and off medication in Parkinson’s disease. Brain Cogn 68, 30–41 (2008).

13. Lewis, M. M. et al. Task specific influences of Parkinson’s disease on the striato-thalamo-cortical and cerebello-thalamo-cortical motor circuitries. Neuroscience 147, 224–235 (2007).

14. Klaus, A., Alves Da Silva, J. & Costa, R. M. What, If, and When to Move: Basal Ganglia Circuits and Self-Paced Action Initiation. Annu Rev Neurosci 42, 459–483 (2019).

15. Baudrexel, S. et al. Resting state fMRI reveals increased subthalamic nucleus–motor cortex connectivity in Parkinson’s disease. Neuroimage 55, 1728–1738 (2011).

16. Nambu, A., Tokuno, H. & Takada, M. Functional significance of the cortico-subthalamo-pallidal ‘hyperdirect’ pathway. Neurosci Res 43, 111–117 (2002).

17. Bonnevie, T. & Zaghloul, K. A. The Subthalamic Nucleus: Unravelling New Roles and Mechanisms in the Control of Action. Neuroscientist 25, 48 (2018).

18. Da Silva, J. A., Tecuapetla, F., Paixão, V. & Costa, R. M. Dopamine neuron activity before action initiation gates and invigorates future movements. Nature 2018 554:7691 554, 244–248 (2018).

19. Ghai, S., Ghai, I., Schmitz, G. & Effenberg, A. O. Effect of rhythmic auditory cueing on parkinsonian gait: A systematic review and meta-analysis. Scientific Reports 2018 8:1 8, 1–19 (2018).

20. Thaut, M. H. et al. Rhythmic auditory stimulation in gait training for Parkinson’s disease patients. Movement Disorders 11, 193–200 (1996).

21. Naro, A., Pignolo, L., Bruschetta, D. & Calabrò, R. S. What about the role of the cerebellum in music-associated functional recovery? A secondary EEG analysis of a randomized clinical trial in patients with Parkinson disease. Parkinsonism Relat Disord 96, 57–64 (2022).

22. Thaut, M. H. Entrainment and the motor system. Music Ther Perspect 31, 31–34 (2013).

23. Kotz, S. A. E. & Schwartze, M. Differential input of the supplementary motor area to a dedicated temporal processing network: Functional and clinical implications. Front Integr Neurosci (2011) doi:10.3389/fnint.2011.00086.

24. Herz, D. M. et al. Dynamic control of decision and movement speed in the human basal ganglia. Nature Communications 2022 13:1 13, 1–15 (2022).

25. Kameda, M., Niikawa, K., Uematsu, A. & Tanaka, M. Sensory and motor representations of internalized rhythms in the cerebellum and basal ganglia. Proc Natl Acad Sci U S A 120, e2221641120 (2023).

26. Buhusi, C. V. & Meck, W. H. What makes us tick? Functional and neural mechanisms of interval timing. Nature Reviews Neuroscience Preprint at 10.1038/nrn1764 (2005).

27. Huang, C. S. et al. Conveyance of cortical pacing for parkinsonian tremor-like hyperkinetic behavior by subthalamic dysrhythmia. Cell Rep 35, 109007 (2021).

28. Levy, R., Hutchison, W. D., Lozano, A. M. & Dostrovsky, J. O. High-frequency Synchronization of Neuronal Activity in the Subthalamic Nucleus of Parkinsonian Patients with Limb Tremor. Journal of Neuroscience 20, 7766–7775 (2000).

29. Hammond, C., Bergman, H. & Brown, P. Pathological synchronization in Parkinson’s disease: networks, models and treatments. Trends Neurosci 30, 357–364 (2007).

30. Chikermane, M. et al. Cortical beta oscillations map to shared brain networks modulated by dopamine. Elife 13, (2024).

31. Brown, P. et al. Dopamine Dependency of Oscillations between Subthalamic Nucleus and Pallidum in Parkinson’s Disease. Journal of Neuroscience 21, 1033–1038 (2001).

32. Gao, L. L., Zhang, J. R., Chan, P. & Wu, T. Levodopa Effect on Basal Ganglia Motor Circuit in Parkinson’s Disease. CNS Neurosci Ther 23, 76–86 (2017).

33. Sharott, A. et al. Spatio-temporal dynamics of cortical drive to human subthalamic nucleus neurons in Parkinson’s disease. Neurobiol Dis 112, 49–62 (2018).

34. Cassidy, M. et al. Movement related changes in synchronization in the human basal ganglia. Brain 125, 1235–1246 (2002).

35. Jenkinson, N. & Brown, P. New insights into the relationship between dopamine, beta oscillations and motor function. Trends Neurosci 34, 611–618 (2011).

36. Baarbé, J. et al. Cortical modulations before lower limb motor blocks are associated with freezing of gait in Parkinson’s disease: an EEG source localization study. Neurobiol Dis 199, (2024).

37. Gulberti, A. et al. Premotor cortical beta synchronization and the network neuromodulation of externally paced finger tapping in Parkinson’s disease. Neurobiol Dis 197, (2024).

38. Yeh, C. H. et al. Auditory cues modulate the short timescale dynamics of STN activity during stepping in Parkinson’s disease. Brain Stimul 17, 501–509 (2024).

39. Williams, D. et al. The relationship between oscillatory activity and motor reaction time in the parkinsonian subthalamic nucleus. European Journal of Neuroscience 21, 249–258 (2005).

40. Praamstra, P. & Pope, P. Slow brain potential and oscillatory EEG manifestations of impaired temporal preparation in Parkinson’s disease. J Neurophysiol 98, 2848–2857 (2007).

41. Te Woerd, E. S., Oostenveld, R., De Lange, F. P. & Praamstra, P. Impaired auditory-to-motor entrainment in Parkinson’s disease. J Neu-rophysiol 117, 1853–1864 (2017).

42. Solís-Vivanco, R., Rodríguez-Violante, M., Cervantes-Arriaga, A., Justo-Guillén, E. & Ricardo-Garcell, J. Brain oscillations reveal impaired novelty detection from early stages of Parkinson’s disease. Neuroimage Clin 18, 923 (2018).

43. Litvak, V. et al. Resting oscillatory cortico-subthalamic connectivity in patients with Parkinson’s disease. Brain (2011) doi:10.1093/brain/awq332.

44. Lee, L. H. N. et al. An electrophysiological perspective on Parkinson’s disease: symptomatic pathogenesis and therapeutic approaches. Journal of Biomedical Science 2021 28:1 28, 1–14 (2021).

45. Engel, A. K. & Fries, P. Beta-band oscillations-signalling the status quo? Current Opinion in Neurobiology Preprint at 10.1016/j.conb.2010.02.015 (2010).

46. Arnal, L. H. Predicting “When” Using the Motor System’s Beta-Band Oscillations. Front Hum Neurosci 6, (2012).

47. Abbasi, O. & Gross, J. Beta-band oscillations play an essential role in motor–auditory interactions. Hum Brain Mapp 41, 656–665 (2020).

48. Fujioka, T., Trainor, L. J., Large, E. W. & Ross, B. Internalized Timing of Isochronous Sounds Is Represented in Neuromagnetic Beta Oscillations. Journal of Neuroscience 32, 1791–1802 (2012).

49. Kühn, A. A. et al. Pathological synchronisation in the subthalamic nucleus of patients with Parkinson’s disease relates to both bradykinesia and rigidity. Exp Neurol 215, 380– 387 (2009).

50. Kühn, A. A., Kupsch, A., Schneider, G. H. & Brown, P. Reduction in subthalamic 8-35 Hz oscillatory activity correlates with clinical improvement in Parkinson’s disease. European Journal of Neuroscience 23, 1956–1960 (2006).

51. Priori, A. et al. Rhythm-specific pharmacological modulation of subthalamic activity in Parkinson’s disease. Exp Neurol 189, 369–379 (2004).

52. Little, S. et al. Bilateral functional connectivity of the basal ganglia in patients with Parkinson’s disease and its modulation by dopaminergic treatment. PLoS One 8, (2013).

53. Cao, C. et al. L-dopa treatment increases oscillatory power in the motor cortex of Parkinson’s disease patients. Neuroimage Clin 26, (2020).

54. Jahanshahi, M. et al. Dopaminergic modulation of striato-frontal connectivity during motor timing in Parkinson’s disease. Brain 133, 727–745 (2010).

55. Evangelisti, S. et al. L-dopa modulation of brain connectivity in Parkinson’s disease patients: A pilot EEG-fmri study. Front Neurosci 13, 462352 (2019).

56. Melgari, J. M. et al. Alpha and beta EEG power reflects L-dopa acute administration in parkinsonian patients. Front Aging Neurosci 6, 302 (2014).

57. Kühn, A. A. et al. High-Frequency Stimulation of the Subthalamic Nucleus Suppresses Oscillatory β Activity in Patients with Parkinson’s Disease in Parallel with Improvement in Motor Performance. Journal of Neuroscience 28, 6165–6173 (2008).

58. Kühn, A. A. et al. Event-related beta desynchronization in human subthalamic nucleus correlates with motor performance. Brain 127, 735–746 (2004).

59. Kühn, A. A. et al. Pathological synchronisation in the subthalamic nucleus of patients with Parkinson’s disease relates to both bradykinesia and rigidity. Exp Neurol 215, 380– 387 (2009).

60. López-Azcárate, J. et al. Coupling between Beta and High-Frequency Activity in the Human Subthalamic Nucleus May Be a Pathophysiological Mechanism in Parkinson’s Disease. Journal of Neuroscience 30, 6667–6677 (2010).

61. Chen, C. C. et al. Stimulation of the subthalamic region at 20 Hz slows the development of grip force in Parkinson’s disease. Exp Neurol 231, 91–96 (2011).

62. Tinkhauser, G. et al. The modulatory effect of adaptive deep brain stimulation on beta bursts in Parkinson’s disease. Brain 140, 1053–1067 (2017).

63. Herz, D. M. et al. Dynamic modulation of subthalamic nucleus activity facilitates adaptive behavior. PLoS Biol 21, e3002140 (2023).

64. Howe, T. E., Lövgreen, B., Cody, F. W. J., Ashton, V. J. & Oldham, J. A. Auditory cues can modify the gait of persons with early-stage Parkinson’s disease: A method for enhancing parkinsonian walking performance? Clin Rehabil 17, 363–367 (2003).

65. Brown, L. A., de Bruin, N., Doan, J. B., Suchowersky, O. & Hu, B. Novel Challenges to Gait in Parkinson’s Disease: The Effect of Concurrent Music in Single-and Dual-Task Contexts. Arch Phys Med Rehabil 90, 1578–1583 (2009).

66. Feigin, A. et al. Effects of levodopa on motor sequence learning in Parkinson’s disease. Neurology 60, 1744–1749 (2003).

67. Rothlind, J. C. et al. Neuropsychological changes following deep brain stimulation surgery for Parkinson’s disease: comparisons of treatment at pallidal and subthalamic targets versus best medical therapy. J Neurol Neurosurg Psychiatry 86, 622–629 (2015).

68. Cools, R. & D’Esposito, M. Inverted-U-shaped dopamine actions on human working memory and cognitive control. Biol Psychiatry 69, e113–e125 (2011).

69. Neumann, W. J., Gilron, R., Little, S. & Tinkhauser, G. Adaptive Deep Brain Stimulation: From Experimental Evidence Toward Practical Implementation. Movement Disorders 38, 937–948 (2023).

70. Williams, N. R., Foote, K. D. & Okun, M. S. Subthalamic Nucleus Versus Globus Pallidus Internus Deep Brain Stimulation: Translating the Rematch Into Clinical Practice. Mov Disord Clin Pract 1, 24–35 (2014).

71. Ferrazzoli, D. et al. Basal ganglia and beyond: The interplay between motor and cognitive aspects in Parkinson’s disease rehabilitation. Neurosci Biobehav Rev 90, 294–308 (2018).

72. Fahn, S., Elton, R. L. & UPDRS program members. Unified Parkinson’s Disease Rating Scale in Recent Developments in Parkinsons Disease. (Macmillan Healthcare Information, 1987).

73. Oostenveld, R., Fries, P., Maris, E. & Schoffelen, J. M. FieldTrip: Open source software for advanced analysis of MEG, EEG, and invasive electrophysiological data. Comput Intell Neurosci (2011) doi:10.1155/2011/156869.

74. Criscuolo, A., Schwartze, M., Bonetti, L. & Kotz, S. A. Aging Impacts Basic Auditory and Timing Processes. European Journal of Neuroscience 61, e70031 (2025).

75. Criscuolo, A., Schwartze, M., Nozaradan, S. & Kotz, S. A. Basal ganglia and cerebellar lesions causally impact the neural encoding of temporal regularities. Imaging Neuroscience 3, (2025).

76. Criscuolo, A., Schwartze, M., Henry, M. J., Obermeier, C. & Kotz, S. A. Individual neurophysiological signatures of spontaneous rhythm processing. Neuroimage 273, 120090 (2023).

77. Criscuolo, A. et al. Macaque monkeys and humans sample temporal regularities in the acoustic environment. Prog Neurobiol 229, 102502 (2023).

78. Cohen, M. X. Analyzing Neural Time Series Data: Theory and Practice. MIT Press Preprint at 10.1017/CBO9781107415324.004 (2014).

79. Voytek, B. et al. Age-Related Changes in 1/f Neural Electrophysiological Noise. Journal of Neuroscience 35, 13257–13265 (2015).

80. Robertson, M. M. et al. EEG power spectral slope differs by ADHD status and stimulant medication exposure in early childhood. J Neurophysiol 122, 2427–2437 (2019).

81. Donoghue, T. et al. Parameterizing neural power spectra into periodic and aperiodic components. Nature Neuroscience 2020 23:12 23, 1655–1665 (2020).

82. Nozaradan, S., Schönwiesner, M., Keller, P. E., Lenc, T. & Lehmann, A. Neural bases of rhythmic entrainment in humans: critical transformation between cortical and lower-level representations of auditory rhythm. European Journal of Neuroscience (2018) doi:10.1111/ejn.13826.

83. Luo, Y. et al. Global, regional, national epidemiology and trends of Parkinson’s disease from 1990 to 2021: findings from the Global Burden of Disease Study 2021. Front Aging Neurosci 16, 1498756 (2025).

84. Morris, M. E., Huxham, F., McGinley, J., Dodd, K. & Iansek, R. The biomechanics and motor control of gait in Parkinson disease. Clinical Biomechanics 16, 459–470 (2001).

85. Dick, J. P. R. et al. The Bereitschaftspotential, L-DOPA and Parkinson’s disease. Electroencephalogr Clin Neurophysiol 66, 263–274 (1987).

86. Dick, J. P. R. et al. The Bereitschaftspotential is abnormal in Parkinson’s disease. Brain 112 (Pt 1), 233–244 (1989).

87. Schurger, A., Hu, P. Ben, Pak, J. & Roskies, A. L. What Is the Readiness Potential? Trends Cogn Sci 25, 558–570 (2021).

88. Grahn, J. A. The role of the basal ganglia in beat perception: Neuroimaging and neuropsychological investigations. in Annals of the New York Academy of Sciences vol. 1169 35–45 (2009).

89. Bartolo, R., Prado, L. & Merchant, H. Information Processing in the Primate Basal Ganglia during Sensory-Guided and Internally Driven Rhythmic Tapping. Journal of Neuroscience 34, 3910–3923 (2014).

90. Merchant, H., Luciana, M., Hooper, C., Majestic, S. & Tuite, P. Interval timing and Parkinson’s disease: Heterogeneity in temporal performance. Exp Brain Res 184, 233–248 (2008).

91. Bareš, M., Lungu, O. V., Husárová, I. & Gescheidt, T. Predictive motor timing performance dissociates between early diseases of the cerebellum and parkinson’s disease. Cerebellum 9, 124–135 (2010).

92. Jones, C. R. G. et al. Modeling accuracy and variability of motor timing in treated and untreated Parkinson’s disease and healthy controls. Front Integr Neurosci 5, 14040 (2011).

93. Claassen, D. O. et al. Deciphering the impact of cerebellar and basal ganglia dysfunction in accuracy and variability of motor timing. Neuropsychologia 51, 267–274 (2013).

94. Grahn, J. A. & Watson, S. L. Perspectives on Rhythm Processing In Motor Regions of the Brain. Music Ther Perspect 31, 25–30 (2013).

95. Grahn, J. A. & Rowe, J. B. Finding and feeling the musical beat: Striatal dissociations between detection and prediction of regularity. Cerebral Cortex (2013) doi:10.1093/cercor/bhs083.

96. Tomassini, A., Ruge, D., Galea, J. M., Penny, W. & Bestmann, S. The Role of Dopamine in Temporal Uncertainty. J Cogn Neurosci 28, 96–110 (2016).

97. Schwartze, M. & Kotz, S. A. Timing Patterns in the Extended Basal Ganglia System. Adv Exp Med Biol 1455, 275–282 (2024).

98. Petter, E. A., Lusk, N. A., Hesslow, G. & Meck, W. H. Interactive roles of the cerebellum and striatum in sub-second and supra-second timing: Support for an initiation, continuation, adjustment, and termination (ICAT) model of temporal processing. Neurosci Biobehav Rev 71, 739–755 (2016).

99. Nougaret, S. et al. Low and high beta rhythms have different motor cortical sources and distinct roles in movement control and spatiotemporal attention. PLoS Biol 22, e3002670 (2024).

100. Cao, C. et al. Low-beta versus high-beta band cortico-subcortical coherence in movement inhibition and expectation. Neurobiol Dis 201, 106689 (2024).

101. Kühn, A. A. et al. Frequency-specific effects of stimulation of the subthalamic area in treated Parkinson’s disease patients. Neuroreport 20, 975–978 (2009).

102. Gulberti, A. et al. Subthalamic deep brain stimulation improves auditory sensory gating deficit in Parkinson’s disease. Clinical Neurophysiology 126, 565–574 (2015).

103. Airaksinen, K. et al. Effects of DBS on auditory and somatosensory processing in Parkinson’s disease. Hum Brain Mapp 32, 1091–1099 (2011).

104. Alain, C., Zendel, B. R., Hutka, S. & Bidelman, G. M. Turning down the noise: the benefit of musical training on the aging auditory brain. Hear Res 308, 162–173 (2014).

105. Bidelman, G. M., Villafuerte, J. W., Moreno, S. & Alain, C. Age-related changes in the subcortical–cortical encoding and categorical perception of speech. Neurobiol Aging 35, 2526–2540 (2014).

106. Brinkmann, P. et al. About time: Ageing influences neural markers of temporal predictability. Biol Psychol 163, 108135 (2021).

107. Haumann, N. T., Petersen, B., Vuust, P. & Brattico, E. Age differences in central auditory system responses to naturalistic music. Biol Psychol 179, 108566 (2023).

108. Herrmann, B., Henry, M. J., Johnsrude, I. S. & Obleser, J. Altered temporal dynamics of neural adaptation in the aging human auditory cortex. Neurobiol Aging 45, 10–22 (2016).

109. Ruohonen, E. M. et al. Event-Related Potentials to Changes in Sound Intensity Demonstrate Alterations in Brain Function Related to Depression and Aging. Front Hum Neurosci 14, (2020).

110. Bonetti, L. et al. Age-related neural changes underlying long-term recognition of musical sequences. Communications Biology 2024 7:1 7, 1–18 (2024).

111. S Leung, A. W., et al. Age Differences in the Neuroelectric Adaptation to Meaningful Sounds. PLoS One 8, 68892 (2013).

112. Voytek, B. et al. Age-Related Changes in 1/f Neural Electrophysiological Noise. Journal of Neuroscience 35, 13257–13265 (2015).

113. Wiest, C. et al. The aperiodic exponent of subthalamic field potentials reflects excitation/inhibition balance in Parkinsonism. Elife 12, (2023).

114. Lang, A. E. & Obeso, J. A. Challenges in Parkinson’s disease: restoration of the nigrostriatal dopamine system is not enough. Lancet Neurol 3, 309–316 (2004).

115. Yıldırım, E. et al. Lower oddball event-related EEG delta and theta responses in patients with dementia due to Parkinson’s and Lewy body than Alzheimer’s disease. Neurobiol Aging 137, 78–93 (2024).

